# Treatment Adherence and Health-Related Quality of Life Among Patients with Type 2 Diabetes Mellitus in Rajshahi, Bangladesh: A Cross-Sectional Study

**DOI:** 10.64898/2026.09.09.26362693

**Authors:** Md. Shahinur Rahaman, Md Rizwanul Karim, Subrata Kumer Sen, A.B.M. Nafiz Intekhab, Suraiya Iasmin, Afroza Akter

## Abstract

**Background:** Type 2 diabetes mellitus (T2DM) is a rapidly growing public health crisis in Bangladesh, where an estimated 13.1 million adults were living with the condition in 2021, with projections suggesting this figure will nearly 22.3 million by 2045. Poor medication adherence and impaired health-related quality of life (HRQoL) are recognized consequences of long-term T2DM management, yet data from northwestern Bangladesh remain scarce. This study aimed to assess treatment adherence, HRQoL and identify their associated factors among T2DM patients in Rajshahi, Bangladesh.

**Methods:** We conducted a cross-sectional study between January and December 2021 among 205 T2DM patients from two tertiary-level hospitals in Rajshahi. Treatment adherence was assessed using the eight-item Morisky Medication Adherence Scale (MMAS-8), and HRQoL was measured using the EuroQol-5 Dimensions Questionnaire (EQ-5D-5L).

**Results:** The mean age of participants was 54.43 (±10.75) years; 50.73% were female. The majority (85.85%) had HbA1c ≥6.5%, reflecting widespread poor glycaemic control. Regarding treatment adherence, 40.98% of patients had low adherence, 26.83% had medium adherence, and 32.19% had high adherence. In terms of HRQoL, only 6.83% were in perfect health state, while 52.68% had slight/moderate problems and 40.49% had severe/extreme problems across EQ-5D-5L domains. The association between treatment adherence and HRQoL was not statistically significant (p=0.265). Multiple regression analysis (Adjusted R²=0.620) identified diabetic complications, illiteracy, primary education, female gender, higher BMI, joint family type, treatment adherence, older age, and two antidiabetic medications as significant predictors of poorer HRQoL, while urban residence and family history of DM were independently associated with better HRQoL.

**Conclusions:** A substantial proportion of T2DM patients exhibited low treatment adherence and markedly impaired HRQoL. While adherence and HRQoL were not directly associated, diabetic complications emerged as the strongest independent predictor of poor quality of life. These findings highlight the urgent need for multidisciplinary interventions to prevent complications in this population.

## 1. BACKGROUND

Diabetes mellitus (DM) is a chronic metabolic disorder characterised by persistent hyperglycaemia resulting from impaired insulin secretion, insulin action, or both, leading to progressive derangements in carbohydrate, lipid, and protein metabolism [1]. The most prevalent form, type 2 DM (T2DM), accounts for approximately 90% of all diabetes cases globally and results from a complex interplay of genetic, environmental, and behavioural risk factors [1, 2]. According to the 11th Edition of the International Diabetes Federation (IDF) Diabetes Atlas, an estimated 589 million adults were living with diabetes worldwide in 2024, projected to rise to 853 million by 2050 [3]. The largest relative increase in burden is expected in low- and middle-income countries (LMICs), where approximately 80% of all people with diabetes reside [3].

Bangladesh, a densely populated South Asian LMIC, faces a particularly alarming diabetes burden. In 2024, an estimated 13.9 million Bangladeshi adults were living with diabetes, projected to nearly double to 23.1 million by 2050 [3]. A recent nationally representative survey found the age-standardised prevalence of diabetes among adults to be 9.2% (95% CI: 8.7–9.7), with 61.5% of those affected unaware of their condition [4]. The prevalence of non-communicable diseases, including T2DM, is rising rapidly in Bangladesh, yet the health system remains underprepared to effectively prevent and manage these conditions [5, 6].

Effective pharmacological management is central to T2DM control. However, therapeutic benefits are contingent on adequate medication adherence. The World Health Organization (WHO) defines adherence as the degree to which a patient’s behavior, taking medication, following a diet, and executing lifestyle changes, corresponds with agreed recommendations from a health care provider [7]. In LMICs, average adherence to long-term therapy for chronic diseases is estimated at only 50%, and substantially lower in resource-limited settings [7]. Non-adherence to antidiabetic therapy compromises glycaemic control, increases the risk of microvascular and macrovascular complications, and elevates mortality [8]. Global estimates suggest approximately 38% of people with T2DM do not take their medications as prescribed [9], with rates of low adherence in Bangladesh ranging from 33% to 46% in recent studies [10, 11].

Health-related quality of life (HRQoL) is an especially important outcome in T2DM, given the chronic nature of the disease and the substantial burden of daily self-management [12, 13]. HRQoL encompasses the physical, psychological, and social dimensions of a patient’s wellbeing as influenced by their disease and treatment [14]. In people with T2DM, disease-related complications, diabetes duration, comorbidities, medication adherence, self-management, and psychosocial factors have been associated with HRQoL [13, 14]. Despite the escalating diabetes burden in Bangladesh, published data on treatment adherence, HRQoL, and factors associated with both treatment adherence and HRQoL are very limited, particularly outside Dhaka and in northwestern Bangladesh. Accordingly, this study aimed to assess treatment adherence, HRQoL and identify factors associated with both treatment adherence and HRQoL among patients with T2DM.

## 2. METHODS

### 2.1 Study Design and Setting

We conducted a hospital based cross-sectional study between January and December 2021. Data were collected from both the inpatient wards and outpatient departments of two major tertiary-level referral hospitals in Rajshahi city: Rajshahi Medical College Hospital (RMCH), the largest public teaching hospital in northwestern Bangladesh, and Rajshahi Diabetic Association General Hospital, a specialised diabetes care facility.

### 2.2 Study Population and Eligibility Criteria

The target population comprised patients diagnosed with T2DM attending the inpatient wards and outpatient departments of the two study hospitals. Patients were eligible for inclusion if they were aged ≥25 years, had a confirmed diagnosis of T2DM for at least one year, and provided written informed consent prior to participation in this study. Patients were excluded if they were aged <25 years, had been diagnosed with DM for less than one year, had a diagnosis of type 1 DM or gestational diabetes, and those who declined to consent for participation.

### 2.3 Sample Size and Sampling

The sample size was calculated using the formula n = Z²σ²/e², where Z=1.96 (95% confidence level), σ=0.27 (standard deviation of the EQ-5D utility index from Redekop et al. [15], used in the absence of Bangladeshi reference data at the time of study design), and e=0.037 (5% of the reference mean utility score of 0.74), yielding n=205. Purposive sampling was used to recruit eligible patients attending the study hospitals during the data collection period until the required sample was achieved.

### 2.4 Data Collection Instruments

Sociodemographic data were collected through a structured questionnaire. Clinical and biochemical data, including fasting blood glucose (FBS), 2-hours-after-breakfast blood glucose (2HABF), and glycated haemoglobin (HbA1c) were collected from each participant’s diabetes record book, a standardised document routinely maintained at both study facilities. The most recent values were recorded from the diabetes record book during routine clinical follow-up of the participants at the facilities were used for this study.

#### 2.4.1 Treatment Adherence (MMAS-8)

Treatment adherence to antidiabetic drugs was assessed using the eight-item Morisky Medication Adherence Scale (MMAS-8) [16, 17]. A validated Bangla-language version of the scale was used under license from the copyright holder (© 2007 Donald E. Morisky). The MMAS-8 consists of seven binary (Yes/No) items and one five-point Likert-scale item. Total MMAS-8 scores range from 0 to 8, where higher scores indicate higher adherence. Based on the total score, patients were categorised as: high adherence (score=8), medium adherence (score 6 to <8), and low adherence (score <6) [18].

#### 2.4.2 Health-Related Quality of Life (EQ-5D-5L)

Health-related quality of life (HRQoL) was assessed using the five-level EuroQol five-dimensional questionnaire (EQ-5D-5L) [19], administered in a validated Bangla version. The instrument comprises five dimensions: mobility, self-care, usual activities, pain/discomfort, and anxiety/depression, each rated on a five-level severity scale (1= no problems to 5= extreme problems/unable to). The responses across the five dimensions were combined to generate a five-digit health-state profile, ranging from 11111 (no problems in any dimension) to 55555 (extreme problems in all dimensions). The health-state profiles were converted into a utility index using the EQ-5D-5L value set for England [20], as a Bangladesh-specific value set was not available at the time of the study. Utility index scores range from -0.285 (states worse than death) to 1 (perfect health). For bivariate analysis, health states were categorised as: perfect health (score 11111; no problems in any domain), slight/moderate health state (problems in some domains, none worse than moderate level), and severe/extreme health state (at least one domain rated as severe or extreme) [21]. For multivariable analysis, the EQ-5D-5L utility index was retained as a continuous outcome measure.

### 2.5 Statistical Analysis

All data were analysed using SPSS version 26.0. Sociodemographic and disease-profile variables were summarised using descriptive statistics. Categorical variables are expressed as frequencies and percentages; continuous variables as means ± standard deviation (SD). Chi-square test was used to assess associations between categorical variables and treatment adherence/HRQoL categories. Multiple linear regression was performed to identify independent predictors of HRQoL (EQ-5D utility index as continuous dependent variable), with assumptions of normality, linearity, homoscedasticity, and absence of multicollinearity verified prior to analysis. A two-tailed p<0.05 was considered statistically significant.

### 2.6 Ethical Considerations

Ethical approval was obtained from the Institutional Review Board and Ethical Review Committee of Rajshahi Medical College, Rajshahi (Approval No. RMC/IRB/2021/27). All participants provided written informed consent prior to enrolment. Participation was voluntary, and participants were assured of data confidentiality and their right to withdraw at any time without consequence.

## 3. RESULTS

### 3.1 Sociodemographic Characteristics

A total of 205 T2DM patients were enrolled. The mean age was 54.43 (±10.75) years; 65.9% were aged ≥50 years. Sex distribution was approximately equal (50.73% female, 49.27% male). The majority were Muslim (93.66%), married (94.15%), and from rural areas (51.71%). Regarding education, 42.93% had completed secondary/higher secondary schooling, while 12.68% were illiterate. The majority (69.27%) had an average monthly household income below BDT 15,000. Mean BMI was 24.73 (±3.36) kg/m²; 36.1% were overweight, and 8.3% were obese. Detailed sociodemographic data are presented in Table 1.

**Table 1.** Sociodemographic characteristics of respondents (n=205)

| Variable | Frequency (n) | Percentage (%) |
| --- | --- | --- |
| **Age (years)** |  |  |
| <50 | 70 | 34.1 |
| $\geq 50$ | 135 | 65.9 |
| **Mean ± SD (years)** | 54.43 ± 10.75 | — |
| **Gender** |  |  |
| Male | 101 | 49.3 |
| Female | 104 | 50.7 |
| **Residence** |  |  |
| Rural | 106 | 51.7 |
| Urban | 99 | 48.3 |
| **Religion** |  |  |
| Islam | 192 | 93.6 |
| Hinduism | 12 | 5.9 |
| Christianity | 1 | 0.5 |
| **Marital status** |  |  |
| Married | 193 | 94.1 |
| Widowed/Divorced | 8 | 3.9 |
| Unmarried | 4 | 2.0 |
| **Family type** |  |  |
| Nuclear | 52 | 25.4 |
| Joint | 71 | 34.6 |
| Three-generation | 82 | 40.0 |
| **Education level** |  |  |
| Illiterate | 26 | 12.7 |
| Primary | 52 | 25.4 |
| Secondary/Higher secondary | 88 | 42.9 |
| Graduate or above | 39 | 19.0 |
| **Occupation** |  |  |
| Housewife | 88 | 42.9 |
| Farmer/Day labour | 46 | 22.4 |
| Businessman | 27 | 13.2 |
| Teacher | 20 | 9.8 |
| Other (banker, engineer, retired, etc.) | 24 | 11.7 |
| **Average monthly income** |  |  |
| <BDT 15,000 | 142 | 69.3 |
| BDT 15,000-25,000 | 27 | 13.2 |
| >BDT 25,000 | 36 | 17.6 |
| Mean ± SD (BDT) | 15,921.95 ± 11,380.38 | — |
| **BMI (kg/m<sup>2</sup>)** |  |  |
| Underweight (<18.5) | 4 | 2.0 |
| Normal (18.5-24.9) | 110 | 53.7 |
| Overweight (25.0-29.9) | 74 | 36.1 |
| Obese (≥30.0) | 17 | 8.3 |
| Mean ± SD | 24.73 ± 3.36 | — |
| **Tobacco use** |  |  |
| No | 146 | 71.2 |
| Yes | 59 | 28.8 |
| – Smoked tobacco | 36 | 61.0* |
| – Smokeless tobacco | 23 | 39.0* |

BDT = Bangladeshi Taka. \*Percentage among tobacco users only.

### 3.2 Disease Profile

The mean duration of T2DM was 10.30 (±6.68) years. The majority (85.85%) had HbA1c ≥6.5% and 90.24% had uncontrolled glycaemic status. More than half (55.12%) had at least one comorbidity, with hypertension being most prevalent (36.6%), followed by hyperlipidaemia (16.6%). A total of 40.49% of respondents had at least one diabetic complication, the most common being acute stroke (15.1%) and nephropathy (13.7%). The vast majority (83.41%) did not engage in regular physical exercise. Disease profile data are presented in Table 2.

**Table 2.** Disease profile of respondents (n=205)

| Variable | Frequency (n) | Percentage (%) |
| --- | --- | --- |
| **Duration of DM** |  |  |
| <10 years | 126 | 61.5 |
| $\geq 10$ years | 79 | 38.5 |
| **Mean ± SD (years)** | 10.30 ± 6.68 | — |
| **HbA1c level** |  |  |
| Normal (<6.0%) | 13 | 6.3 |
| Pre-diabetes (6.0-6.4%) | 16 | 7.8 |
| Diabetes ($\geq 6.5\%$) | 176 | 85.9 |
| **Fasting blood sugar (FBS) level (mmol/L)** |  |  |
| Normal (<6.1) | 16 | 7.8 |
| Impaired fasting glucose (6.1–6.9) | 38 | 18.5 |
| Diabetes ($\geq 7.0$) | 151 | 73.7 |
| **2 Hours After Breakfast (2HABF) blood sugar level (mmol/L)** |  |  |
| Normal (<7.8) | 16 | 7.8 |
| Impaired glucose tolerance (7.8–11.0) | 65 | 31.7 |
| Diabetes ($\geq 11.1$) | 124 | 60.5 |
| **Glycaemic status** |  |  |
| Controlled | 20 | 9.8 |
| Uncontrolled | 185 | 90.2 |
| **Family history of DM** |  |  |
| Yes | 133 | 64.9 |
| No | 72 | 35.1 |
| **Management of DM** |  |  |
| Oral medication only | 91 | 44.4 |
| Insulin injection only | 34 | 16.6 |
| Combination (oral + insulin) | 79 | 38.5 |
| Diet control only | 1 | 0.5 |
| **Number of antidiabetic medications** |  |  |
| One | 39 | 19.0 |
| Two | 79 | 38.5 |
| Three or more | 87 | 42.4 |
| **Comorbidities (any)** |  |  |
| Present | 113 | 55.1 |
| Absent | 92 | 44.9 |
| **Most common comorbidities** |  |  |
| Hypertension | 75 | 36.6 |
| Hyperlipidaemia | 34 | 16.6 |
| Osteoarthritis | 12 | 5.9 |
| Hypothyroidism | 8 | 3.9 |
| Atrial fibrillation | 5 | 2.4 |
| **Diabetic complications (any)** |  |  |
| Present | 83 | 40.5 |
| Absent | 122 | 59.5 |
| **Most common complications** |  |  |
| Acute stroke | 31 | 15.1 |
| Nephropathy | 28 | 13.7 |
| Retinopathy | 8 | 3.9 |
| Foot ulcer | 8 | 3.9 |
| Acute myocardial infarction | 5 | 2.4 |
| Transient ischaemic attack | 4 | 2.0 |
| **Regular physical exercise** |  |  |
| No | 171 | 83.4 |
| Yes | 34 | 16.6 |
| – <150 min/week | 5 | 14.7† |
| – ≥150 min/week | 29 | 85.3† |

### 3.3 Treatment Adherence

Regarding treatment adherence, 84 respondents (40.98%) had low adherence (MMAS-8 score <6), 55 (26.83%) had medium adherence (score 6 to <8), and 66 (32.19%) had high adherence (score 8) (Figure 1).

**Figure 1.**
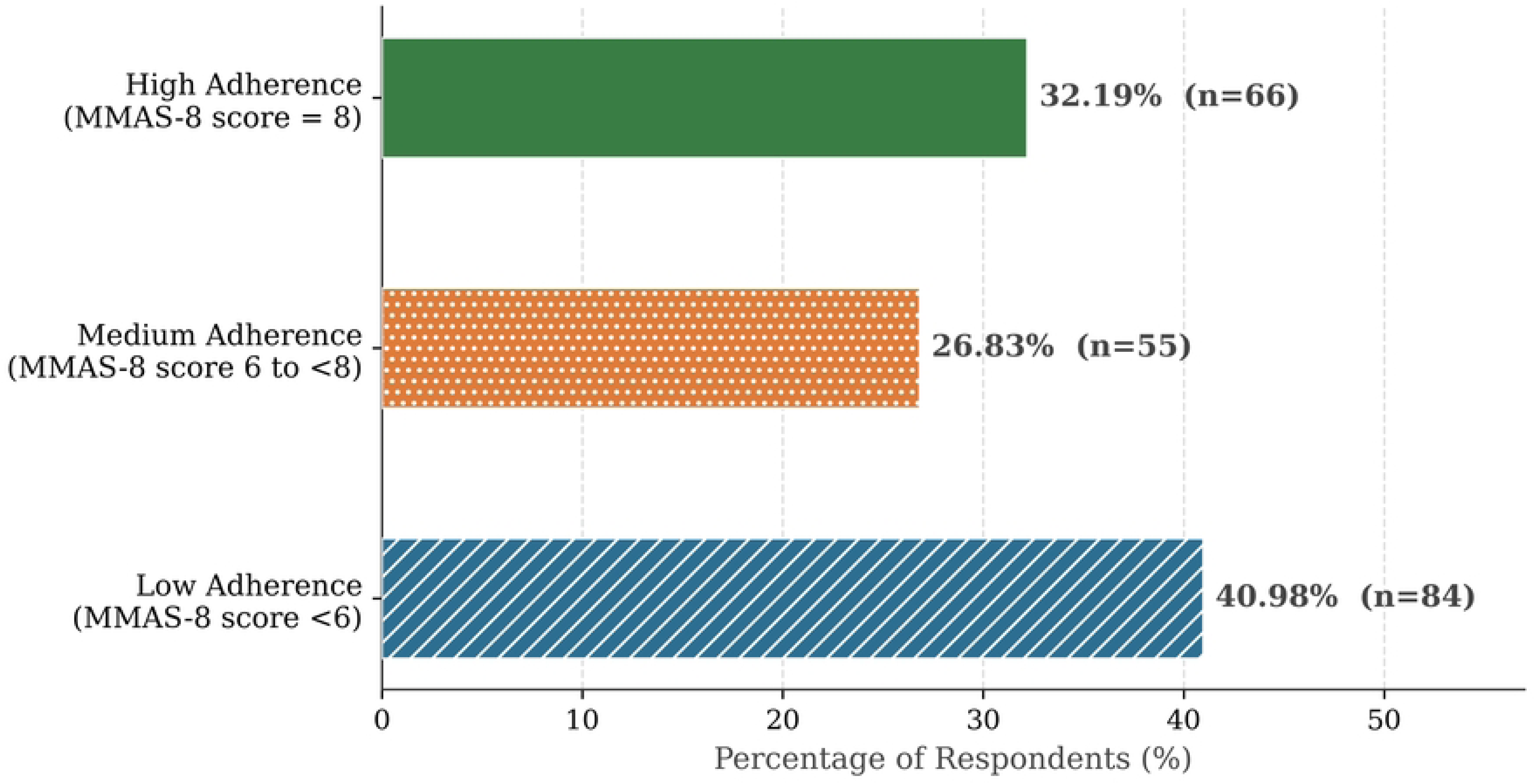
Distribution of respondents by treatment adherence level using MMAS-8 (n=205). Age, sex, residence, education level, average monthly income, BMI, tobacco use, HbA1c level, management modality, comorbidities, and diabetic complications were all significantly associated with treatment adherence level (p<0.05). Duration of DM and family type were not significantly associated with adherence (p>0.05). Notably, tobacco users showed substantially higher rates of low adherence compared to non-users (67.8% vs. 30.1%), and patients with complications had considerably higher rates of low adherence than those without (56.6% vs. 30.3%) (p<0.001) (Table 3).

**Table 3.** Factors associated with treatment adherence level among T2DM patients (n=205)

| Variable | Category | Low adherence<br>n (%) | Medium adherence<br>n (%) | High adherence<br>n (%) | p-value |
| --- | --- | --- | --- | --- | --- |
| **Age** | <50 years (n=70) | 23 (32.9%) | 16 (22.9%) | 31 (44.3%) | 0.028 |
|  | ≥50 years (n=135) | 61 (45.2%) | 39 (28.9%) | 35 (25.9%) |  |
| **Sex** | Male (n=101) | 50 (49.5%) | 22 (21.8%) | 29 (28.7%) | 0.046 |
|  | Female (n=104) | 34 (32.7%) | 33 (31.7%) | 37 (35.6%) |  |
| **Residence** | Rural (n=106) | 50 (47.2%) | 34 (32.1%) | 22 (20.8%) | 0.001 |
|  | Urban (n=99) | 34 (34.3%) | 21 (21.2%) | 44 (44.4%) |  |
| **Education level** | Illiterate (n=26) | 15 (57.7%) | 7 (26.9%) | 4 (15.4%) | 0.001 |
|  | Primary (n=52) | 21 (40.4%) | 18 (34.6%) | 13 (25.0%) |  |
|  | Secondary/HSC (n=88) | 39 (44.3%) | 24 (27.3%) | 25 (28.4%) |  |
|  | Graduate & above (n=39) | 9 (23.1%) | 6 (15.4%) | 24 (61.5%) |  |
| **Average monthly income** | <BDT 15,000 (n=142) | 54 (38.0%) | 45 (31.7%) | 43 (30.3%) | 0.039 |
|  | BDT 15,000-25,000 (n=27) | 10 (37.0%) | 7 (25.9%) | 10 (37.0%) |  |
|  | >BDT 25,000 (n=36) | 20 (55.6%) | 3 (8.3%) | 13 (36.1%) |  |
| **BMI** | Normal/Underweight (n=114) | 39 (34.2%) | 37 (32.4%) | 38 (33.4%) | <0.001 |
|  | Overweight/Obese (n=91) | 45 (49.5%) | 18 (19.7%) | 28 (30.8%) |  |
| **Tobacco use** | No (n=146) | 44 (30.1%) | 43 (29.5%) | 59 (40.4%) | <0.001 |
|  | Yes (n=59) | 40 (67.8%) | 12 (20.3%) | 7 (11.9%) |  |
| **HbA1c level** | <6.0% (n=13) | 4 (30.8%) | 3 (23.1%) | 6 (46.2%) | <0.001 |
|  | 6.0-6.4% (n=16) | 1 (6.3%) | 2 (12.5%) | 13 (81.3%) |  |
|  | ≥6.5% (n=176) | 79 (44.9%) | 50 (28.4%) | 47 (26.7%) |  |
| **DM management** | Oral medication (n=91) | 33 (36.3%) | 23 (25.3%) | 35 (38.5%) | 0.022 |
|  | Insulin injection (n=34) | 15 (44.1%) | 15 (44.1%) | 3 (8.8%) |  |
|  | Combination- oral & insulin (n=79) | 35 (44.3%) | 17 (21.5%) | 27 (34.2%) |  |
| **Comorbidities** | Present (n=113) | 56 (49.6%) | 18 (15.9%) | 39 (34.5%) | <0.001 |
|  | Absent (n=92) | 28 (30.4%) | 37 (40.2%) | 27 (29.4%) |  |
| **Diabetic complications** | Present (n=83) | 47 (56.6%) | 23 (27.7%) | 13 (15.7%) | <0.001 |
|  | Absent (n=122) | 37 (30.3%) | 32 (26.2%) | 53 (43.4%) |  |
| **Duration of DM** | <10 years (n=126) | 46 (36.5%) | 33 (26.2%) | 47 (37.3%) | 0.117 |
|  | ≥10 years (n=79) | 38 (48.1%) | 22 (27.8%) | 19 (24.1%) |  |
| Family type | Nuclear (n=52) | 22 (42.3%) | 11 (21.2%) | 19 (36.5%) | 0.457 |
|  | Joint (n=71) | 25 (35.2%) | 20 (28.2%) | 26 (36.6%) |  |
|  | Three-generation (n=82) | 37 (45.1%) | 24 (29.3%) | 21 (25.6%) |  |

*p-values were calculated using the chi-square ( $\chi^2$ ) test. Statistical significance was considered at $p < 0.05$ .*
*HSC = Higher Secondary Certificate; BDT = Bangladeshi Taka; DM = Diabetes Mellitus.*

### 3.4 Health-Related Quality of Life

Regarding HRQoL, only 14 respondents (6.83%) were in perfect health state (no problems in any EQ-5D-5L domain). The majority, 108 (52.68%), were in a slight/moderate health state, while 83 (40.49%) were in a severe/extreme health state (Figure 2).

**Figure 2.**
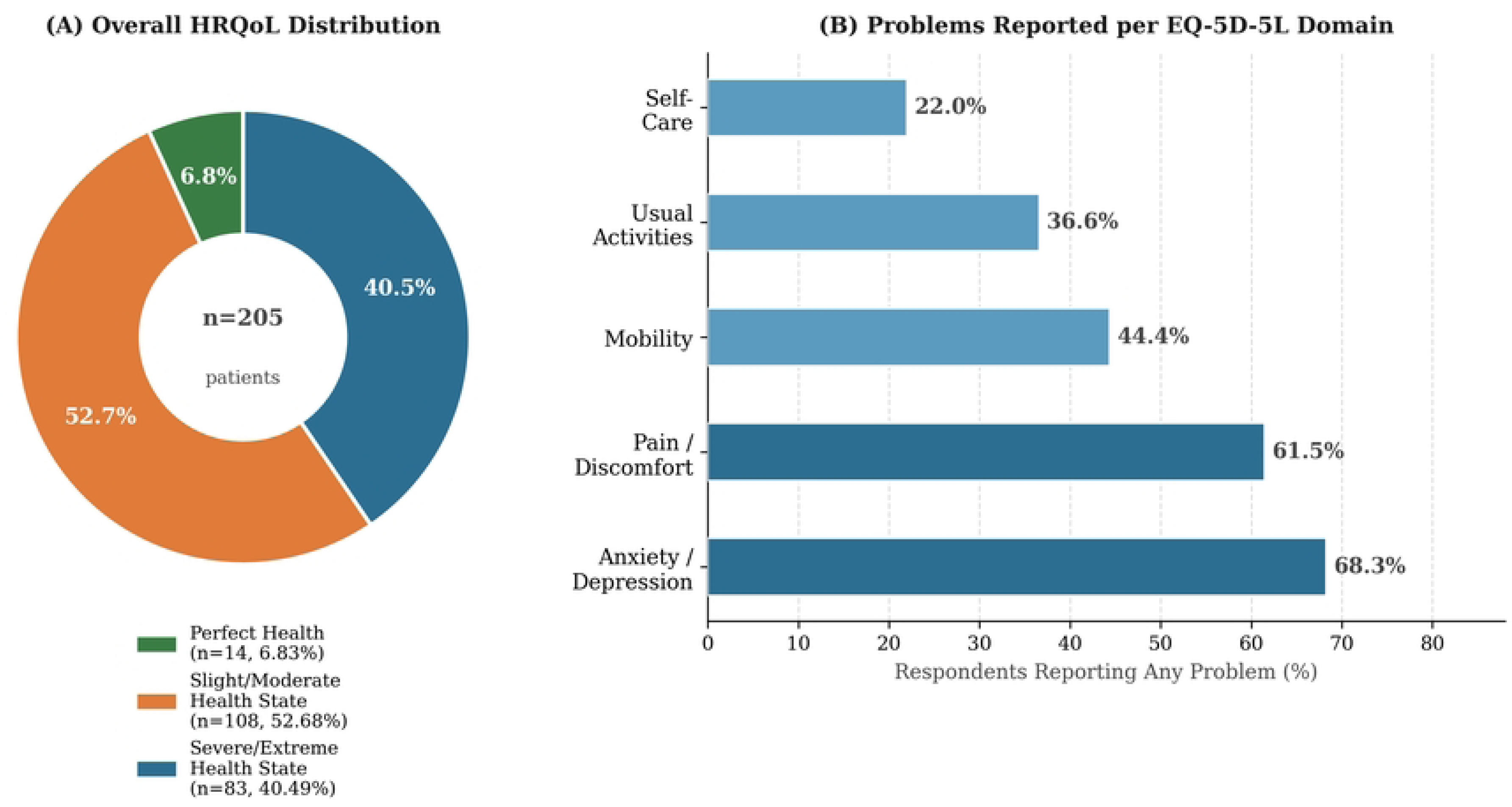
Health-related quality of life (HRQoL) distribution and EQ-5D-5L domain-level problems among T2DM patients in Rajshahi (n=205). Panel A: overall HRQoL distribution; Panel B: proportion reporting any problem per domain. Age, family type, education level, occupation, BMI, HbA1c level, glycaemic status, family history of DM, management modality, number of antidiabetic medications, comorbidities, diabetic complications, and regular physical exercise were all significantly associated with HRQoL (p<0.05). Sex, residence, monthly income, tobacco use, and duration of DM were not significantly associated with HRQoL. Of particular note, among respondents who performed regular physical exercise, none had a severe/extreme health state, compared to 48.5% of non-exercisers (p<0.001). Similarly, among patients with diabetic complications, 71.1% had a severe/extreme health state, compared to only 19.7% of those without complications (p<0.001) (Table 4).

**Table 4.**
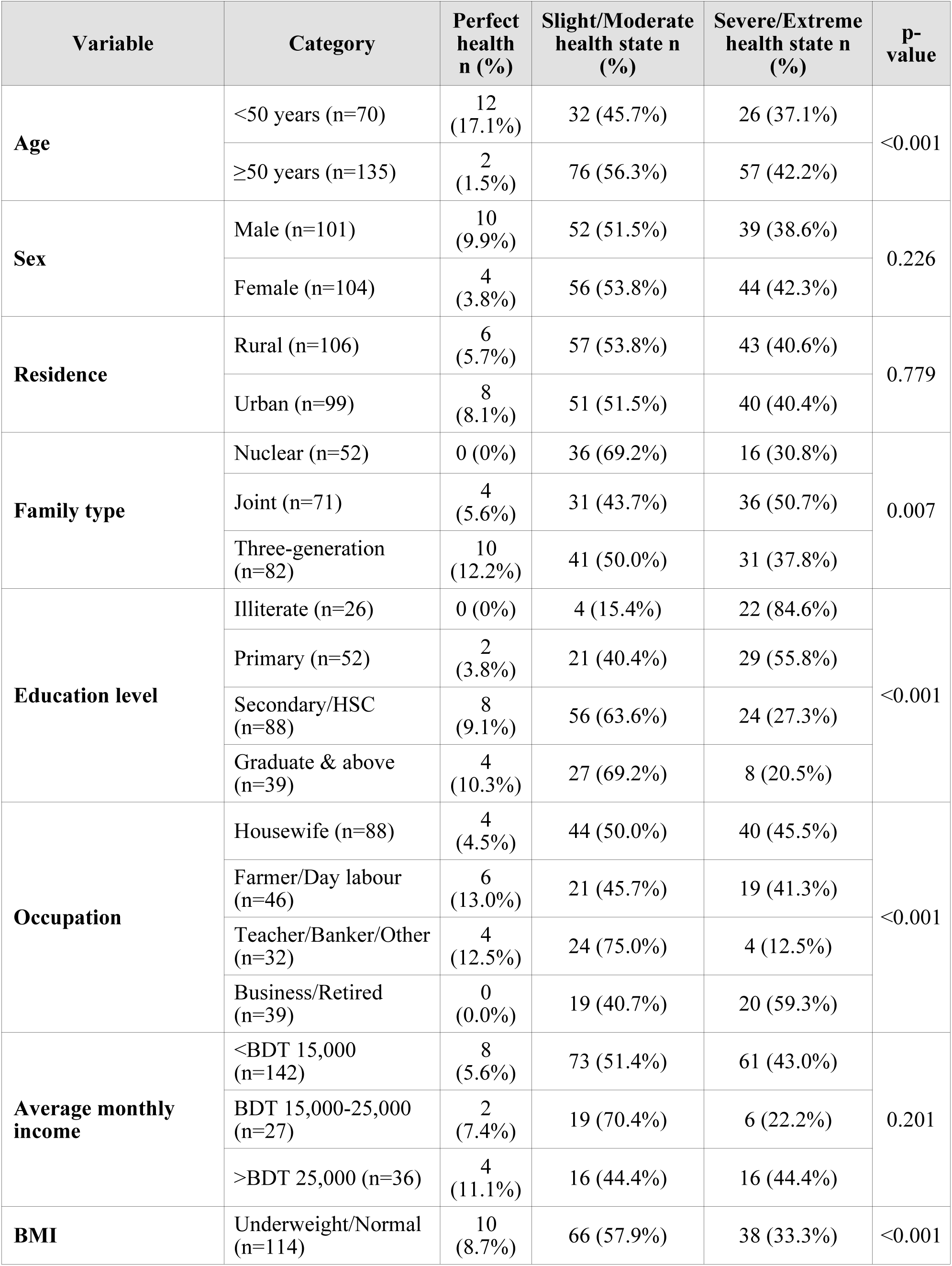
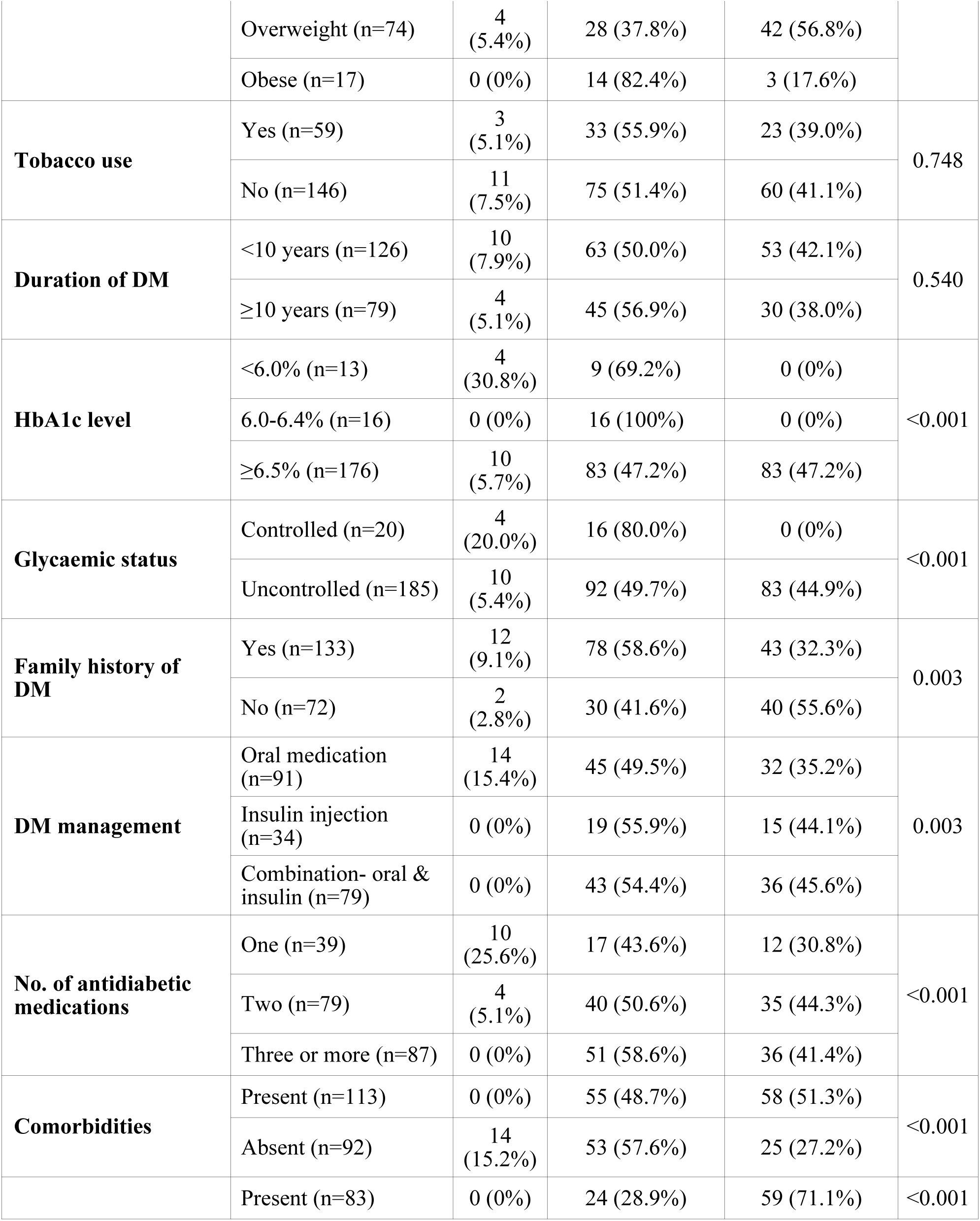
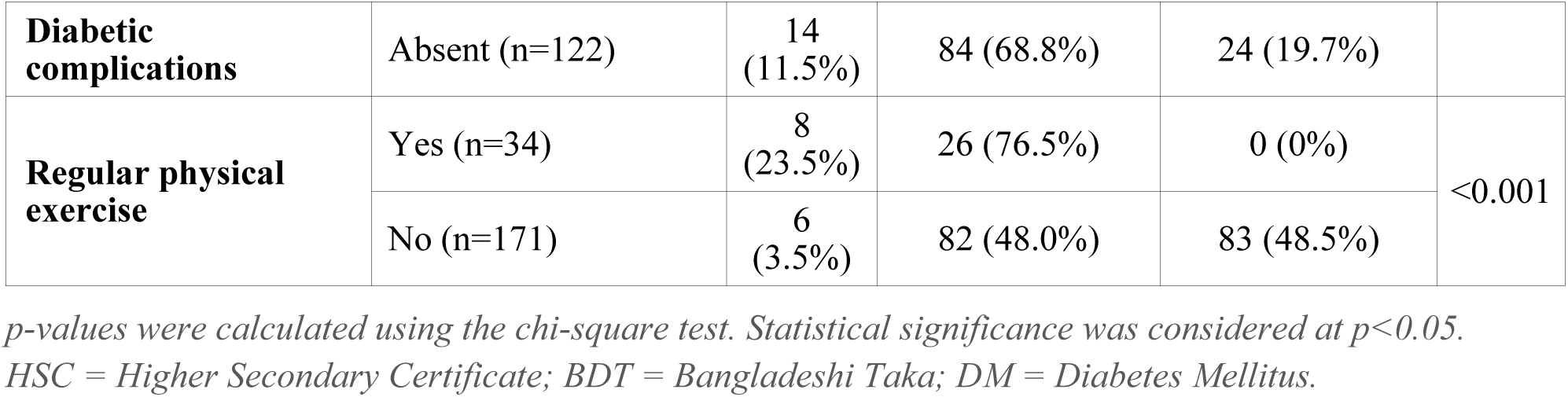
Factors associated with health-related quality of life (HRQoL) among T2DM patients (n=205)

### 3.5 Relationship Between Treatment Adherence and HRQoL

The cross-tabulation of treatment adherence level against EQ-5D-5L health state is presented in Figure 3. Among low adherence patients, 53.6% were in a slight/moderate health state and 44.0% in a severe/extreme health state. Among high adherence patients, 54.5% were in a slight/moderate state and 34.8% in a severe/extreme state. The distribution of HRQoL categories across adherence groups was strikingly similar, and the association was statistically non-significant (p=0.265).

**Figure 3.**
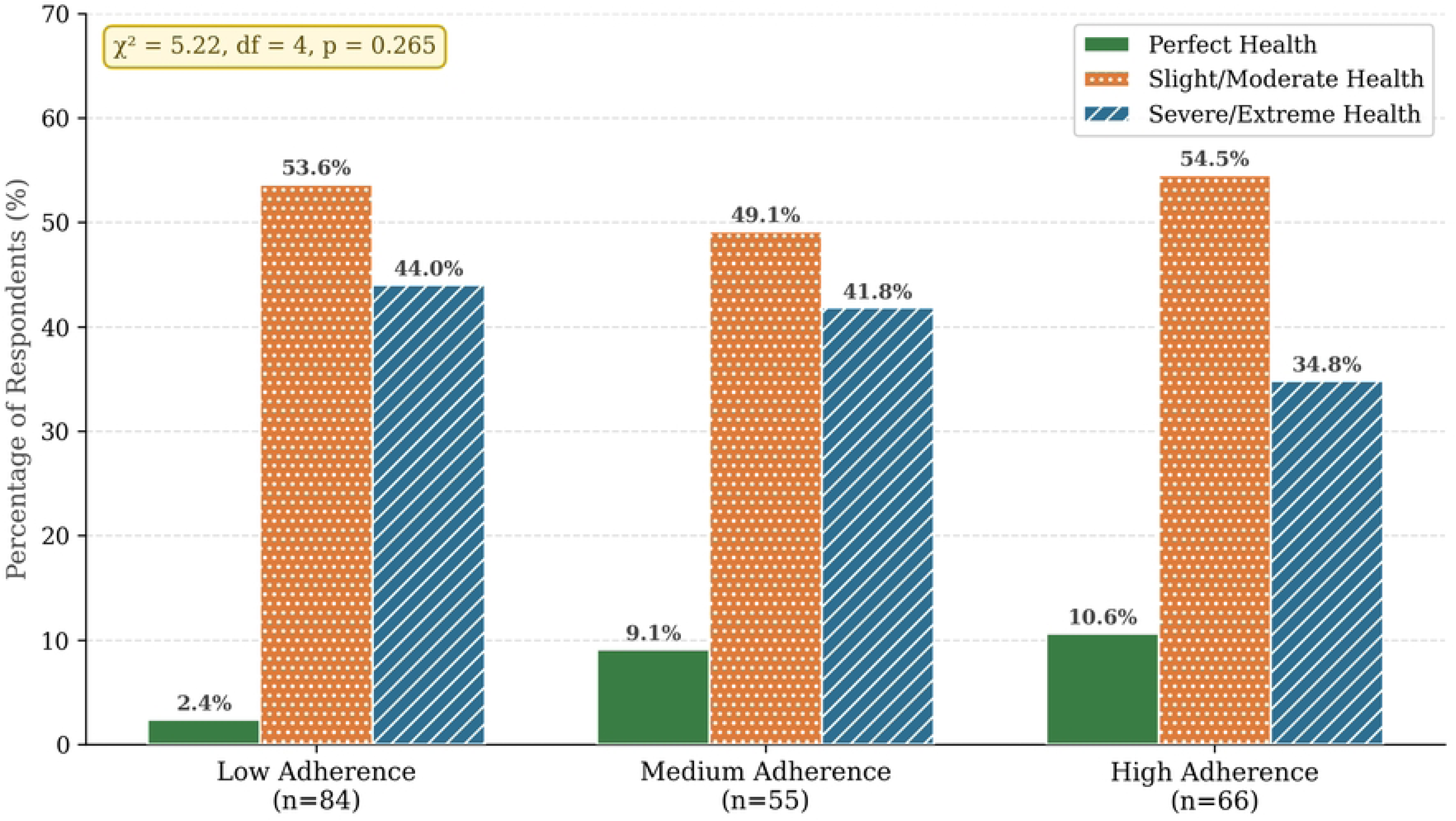
Relationship between treatment adherence level and health-related quality of life category among T2DM patients (n=205).

### 3.6 Predictors of HRQoL: Multiple Regression Analysis

Multiple linear regression was performed with EQ-5D-5L index total score as the dependent variable. The model explained 64.0% of the variance in HRQoL (Adjusted R²=0.620; F(11,193) = 31.240; p<0.001). Independent significant predictors of poorer HRQoL (higher beta associated with worse HRQoL score) were: diabetic complications (β=0.587, p<0.001), illiteracy (β=0.320, p<0.001), primary education level (β=0.206, p<0.001), Female gender (β=0.202, p<0.001), BMI (β=0.196, p<0.001), joint family type (β=0.177, p<0.001), treatment adherence (β=0.177, p<0.001), age (β=0.166, p<0.001), and number of antidiabetic medications, two vs. one (β=0.119, p=0.014). Urban residence (β=−0.153, p=0.010) and family history of DM (β=−0.104, p=0.033) were independently associated with better HRQoL (Table 5).

**Table 5.** Multiple linear regression: independent predictors of HRQoL (EQ-5D-5L utility index) among T2DM patients (n=205)

| Predictor variable | B | $\beta$ | t | p-value | 95% CI for B |
| --- | --- | --- | --- | --- | --- |
| Diabetic complications | 0.383 | 0.587 | 11.634 | <0.001 | (0.318, 0.447) |
| Illiterate (ref: graduate & above) | 0.308 | 0.320 | 6.333 | <0.001 | (0.212, 0.404) |
| Primary education (ref: graduate & above) | 0.152 | 0.206 | 4.127 | <0.001 | (0.079, 0.224) |
| Female gender (ref: male) | 0.129 | 0.202 | 3.912 | <0.001 | (0.064, 0.194) |
| BMI (kg/m<sup>2</sup>) | 0.019 | 0.196 | 3.975 | <0.001 | (0.009, 0.028) |
| Joint family type (ref: nuclear) | 0.119 | 0.177 | 3.798 | <0.001 | (0.034, 0.240) |
| Treatment adherence (MMAS-8 category) | 0.067 | 0.177 | 3.618 | <0.001 | (0.030, 0.103) |
| Age (years) | 0.005 | 0.166 | 3.454 | <0.001 | (0.002, 0.007) |
| Urban residence (ref: rural) | -0.098 | -0.153 | -2.596 | 0.010 | (-0.172, -0.024) |
| Two antidiabetic medications (ref: one) | 0.078 | 0.119 | 2.478 | 0.014 | (0.016, 0.140) |
| Family history of DM (ref: absent) | -0.070 | -0.104 | -2.146 | 0.033 | (-0.134, -0.006) |

Adjusted $R^2 = 0.620$ ; $R^2 = 0.640$ ; $F(11,193) = 31.240$ , $p < 0.001$ .
Predictors ordered by absolute standardised $\beta$ (effect size) descending. B = unstandardised coefficient; $\beta$ = standardised coefficient; CI = confidence interval.

## 4. DISCUSSION

This cross-sectional study evaluated treatment adherence and health-related quality of life (HRQoL) among 205 patients with T2DM attending two tertiary-level hospitals in Rajshahi, Bangladesh. Overall, the largest proportion of patients had low treatment adherence (40.98%).

Only 6.8% of patients reported perfect HRQoL, while 40.49% were in a severe or extreme health state. Although treatment adherence was not significantly associated with HRQoL in the bivariate analysis, multivariable regression identified diabetic complications as the strongest independent predictor of HRQoL, followed by illiteracy, primary education, female gender, higher BMI, joint family type, treatment adherence, older age, two antidiabetic medications, urban residency, and family history of DM.

The prevalence of low treatment adherence (40.98%) is consistent with the broader regional literature. Mannan et al. [10] reported a low adherence prevalence of 46.3% among 2,070 T2DM patients in southern Bangladesh, while Monju et al. [11] found 33.2% low adherence in Chittagong. Globally, approximately 38% of people with T2DM do not adhere to prescribed medications [9]. The somewhat lower non-adherence rate in this study may partly reflect the characteristics of a tertiary-care, specialist-attending population. The finding that 67.8% of tobacco users had low adherence compared to 30.1% of non-users suggests that lifestyle risk behaviours cluster with poor treatment adherence, a clinically important pattern warranting targeted counselling.

In this study, age, sex, residence, education level, average monthly income, BMI, tobacco use, HbA1c level, DM management modality, comorbidities, and diabetic complications were all significantly associated with medication adherence (p<0.05), while duration of DM and family type were not. A cross-sectional study in Saudi Arabia similarly found that medication adherence was significantly associated with age above 50 years, urban residence, HbA1c level, and number of daily medications among T2DM patients [22]. A recent Bangladesh study from rural sub-district health facilities also identified residence as a significant factor associated with medication adherence among diabetic patients [11], corroborating our finding of significantly higher low adherence among rural residents. Regarding tobacco use, a systematic review and meta-analysis of socio-personal factors affecting treatment adherence in T2DM found that the risk of non-adherence was 22% higher in smokers than non-smokers [23], consistent with our finding that 67.8% of tobacco users had low adherence compared to only 30.1% of non-users, suggesting that tobacco use and poor treatment adherence may represent co-occurring health-risk behaviours in this population.

The HRQoL findings are concerning: only 6.83% of patients had perfect health and 40.49% were in a severe or extreme health state. These results are broadly consistent with studies from Bangladesh. Barua et al. [14], in a nationwide cross-sectional study of 1,806 T2DM patients, reported that over half of the participants had average HRQoL and 64% reported problems in at least one EQ-5D-5L domain. A more recent study by Ahmed et al. [24] among 480 diabetic patients in Dhaka using the same instruments found that low medication adherence was associated with the anxiety/depression and pain/discomfort dimensions. The striking finding in this study that none of the physically active patients had a severe/extreme health state, compared to 48.5% of inactive patients (p<0.001), highlights the profound protective role of physical activity on HRQoL, particularly salient in this population where 83.4% were physically inactive.

In this study, age, family type, education level, occupation, BMI, HbA1c level, glycaemic status, family history of DM, DM management modality, number of antidiabetic medications, comorbidities, diabetic complications, and regular physical exercise were all significantly associated with HRQoL (p<0.05), while sex, residence, average monthly income, tobacco use, and duration of DM were not. A systematic review of 35 cross-sectional studies found that older age, lower monthly family income, less physical activity, and the presence of multiple comorbidities were all associated with lower HRQoL in T2DM patients, with age, education, occupation, BMI, HbA1c, treatment modality, complications, and comorbidities consistently identified as significant determinants [13]. Concerning the number of antidiabetic medications, a cross-sectional study among chronically ill patients found that higher medication use was associated with progressively worse EQ-5D-5L utility scores [25], consistent with our finding that patients on two or more antidiabetic medications had significantly poorer HRQoL than those on single-drug regimens. The profound protective effect of regular physical exercise on HRQoL observed in this study, where none of the physically active patients had a severe or extreme health state, compared to 48.5% of inactive patients (p<0.001), is well-supported by evidence. A systematic review and meta-analysis found that aerobic exercise alone (p<0.001) or in combination with resistance training (p<0.001) significantly improved quality of life in adults with T2DM, primarily through improvements in physical components of HRQoL [26]. A large population-based cross-sectional study in Korea using the EQ-5D similarly demonstrated that T2DM patients who engaged in regular daily walking had significantly better HRQoL scores compared to those who were physically inactive [27].

The non-significant association between treatment adherence and HRQoL (p=0.265) is perhaps the most noteworthy finding of this study. While studies by Mishra et al. [28] and Nugraha and Putri [12] reported significant positive associations between adherence and HRQoL, our finding aligns with Martínez et al. [29] and Azmi et al. [30], who found no significant correlation. Multiple explanations may account for this. First, the cross-sectional design precludes inference of temporality: HRQoL reflects cumulative disease progression, while MMAS-8 captures recent adherence behaviour. Second, in this sample, diabetic complications rather than current adherence appear to dominate HRQoL impairment. Third, a recent review in *Diabetologia* reported only weak-to-moderate correlations between MMAS-8 and EQ-5D-5L (rho=0.136-0.230) across T2DM studies [31], supporting the conclusion that this relationship may be modest in magnitude and difficult to detect in cross-sectional samples.

We identified diabetic complications as the single strongest predictor of poor HRQoL (β=+0.587, p<0.001), underscoring the catastrophic impact of complications, particularly stroke and nephropathy, which together affected 28.8% of respondents on patient wellbeing. This finding is consistent with international evidence and reinforces the critical importance of complication prevention as a HRQoL-preserving strategy [13, 14, 21]. Illiteracy and lower educational attainment were independently associated with poorer HRQoL (β=+0.320 and β=+0.206 respectively), plausibly mediated through lower health literacy, suboptimal self-management behaviours, and reduced access to quality care. Female gender was a significant independent predictor of worse HRQoL (β=+0.202, p<0.001), consistent with prior evidence that women with T2DM report greater disease burden, higher rates of anxiety and depression, and lower quality of life compared to men [13]. Higher BMI (β=+0.196, p<0.001) was also a significant predictor, consistent with evidence that overweight and obese T2DM patients experience lower HRQoL than normal-weight patients [13]. The protective effect of urban residence (β=−0.153, p=0.010) may reflect better healthcare infrastructure, access to specialists, and health information availability in urban Rajshahi. Counter-intuitively, family history of DM was associated with better HRQoL (β=−0.104, p=0.033), a finding that may reflect greater disease awareness and preparedness in families with prior DM experience, leading to earlier health-seeking behaviour and better self-management, though this warrants further investigation.

This study has several limitations. First, purposive sampling from two tertiary hospitals limits generalisability to the broader T2DM population; hospital attendees represent a more symptomatic subset. Second, the cross-sectional design prevents causal inference; the direction of any relationship between adherence and HRQoL cannot be established. Third, self-reported adherence via MMAS-8 is susceptible to social desirability bias. Fourth, the England EQ-5D-5L value set was applied in the absence of a validated Bangladeshi value set, which may not accurately reflect local health preferences. Fifth, findings may not represent patients managed in primary care or rural settings.

## 5. CONCLUSIONS

This study demonstrates that low treatment adherence and substantially impaired HRQoL are prevalent among T2DM patients in Rajshahi, Bangladesh. The majority of patients exhibited poor glycaemic control, high complication burden, and physical inactivity, each of which independently predicted worse HRQoL. While treatment adherence and HRQoL were not directly associated at the bivariate level, multivariate analysis revealed that diabetic complications represent the most powerful determinant of quality of life impairment in this population, followed by illiteracy, primary education, female gender, higher BMI, joint family type, treatment adherence, older age, two antidiabetic medications, urban residency, and family history of DM.

These findings have important implications for clinical practice and health policy in Bangladesh. These findings underscore the urgent need for multidisciplinary care interventions integrating physicians, pharmacists, nurses, dietitians, and psychologists for targeting adherence improvement and complication prevention. Regular adherence monitoring, patient counselling, and community-based diabetes education programmes should be incorporated into routine diabetic care, with particular attention to at-risk groups including tobacco users, patients on insulin therapy, and those with established comorbidities. Future longitudinal research and intervention trials are warranted to establish causal relationships and evaluate the effectiveness of adherence-improving strategies on HRQoL outcomes.

## Data Availability

The de-identified dataset generated and analyzed during this study is available from the corresponding author upon reasonable request.

## Acknowledgement

The authors sincerely thank the administration and clinical staff of Rajshahi Medical College Hospital (RMCH) and Rajshahi Diabetic Association General Hospital for their cooperation and support during data collection. We are deeply grateful to all study participants for their time and willingness to share information. The authors also acknowledge the Department of Community Medicine, Rajshahi Medical College, for academic support, and the Morisky Medication Adherence Research and the EuroQol Research Foundation for granting permission to use the MMAS-8 and EQ-5D-5L, respectively.

## REFERENCES

1. David GG DM. Greenspan’s Basic & Clinical Endocrinology. New York: McGraw-Hill Education; 2017.

2. Khan AKA MH. Diabetes Mellitus. Dhaka: Diabetic Association of Bangladesh; 2018.

3. Sun H, Saeedi P, Karuranga S, Pinkepank M, Ogurtsova K, Duncan BB, et al. IDF Diabetes Atlas: Global, regional and country-level diabetes prevalence estimates for 2021 and projections for 2045. Diabetes research and clinical practice. 2022;183:109119.

4. Hossain MB, Khan MN, Oldroyd JC, Rana J, Magliago DJ, Chowdhury EK, et al. Prevalence of, and risk factors for, diabetes and prediabetes in Bangladesh: Evidence from the national survey using a multilevel Poisson regression model with a robust variance. PLOS Global Public Health. 2022;2(6):e0000461.

5. Afroz A, Alam K, Ali L, Karim A, Alramadan MJ, Habib SH, et al. Type 2 diabetes mellitus in Bangladesh: a prevalence based cost-of-illness study. BMC health services research. 2019;19(1):601.

6. Whiting DR, Guariguata L, Weil C, Shaw J. IDF diabetes atlas: global estimates of the prevalence of diabetes for 2011 and 2030. Diabetes research and clinical practice. 2011;94(3):311– 21.

7. Sabaté E. Adherence to long-term therapies: evidence for action: World health organization; 2003.

8. Cramer JA. A systematic review of adherence with medications for diabetes. Diabetes care. 2004;27(5):1218–24.

9. Polonsky WH, Henry RR. Poor medication adherence in type 2 diabetes: recognizing the scope of the problem and its key contributors. Patient preference and adherence. 2016:1299–307.

10. Mannan A, Hasan MM, Akter F, Rana MM, Chowdhury NA, Rawal LB, et al. Factors associated with low adherence to medication among patients with type 2 diabetes at different healthcare facilities in southern Bangladesh. Global health action. 2021;14(1):1872895.

11. Monju IH, Ahmed T, Abrar M, Habib MA, Faiza F, Hawlader MDH, et al. Prevalence and factors associated with medication adherence among diabetes patients in rural sub-district health facilities in Bangladesh. Discover Public Health. 2025;22(1):114.

12. Nugraha S, Putri SK, editors. Adherence and Quality of Life in Patients with Type II Diabetes Mellitus. Social and Humaniora Research Symposium (SoRes 2018); 2019: Atlantis Press.

13. Teli M, Thato R, Rias YA. Predicting factors of health-related quality of life among adults with type 2 diabetes: a systematic review. SAGE open nursing. 2023;9:23779608231185921.

14. Barua L, Faruque M, Chowdhury HA, Banik PC, Ali L. Health-related quality of life and its predictors among the type 2 diabetes population of Bangladesh: A nation-wide cross-sectional study. Journal of diabetes investigation. 2021;12(2):277–85.

15. Redekop WK, Koopmanschap MA, Stolk RP, Rutten GE, Wolffenbuttel BH, Niessen LW. Health-related quality of life and treatment satisfaction in Dutch patients with type 2 diabetes. Diabetes care. 2002;25(3):458–63.

16. Al-Qazaz HK, Hassali MA, Shafie AA, Sulaiman SA, Sundram S, Morisky DE. The eight-item Morisky Medication Adherence Scale MMAS: translation and validation of the Malaysian version. Diabetes research and clinical practice. 2010;90(2):216–21.

17. Krousel-Wood M, Islam T, Webber LS, Re R, Morisky DE, Muntner P. New medication adherence scale versus pharmacy fill rates in hypertensive seniors. The American journal of managed care. 2009;15(1):59.

18. Lee W-Y, Ahn J, Kim J-H, Hong Y-P, Hong SK, Kim YT, et al. Reliability and validity of a self-reported measure of medication adherence in patients with type 2 diabetes mellitus in Korea. Journal of International Medical Research. 2013;41(4):1098–110.

19. Group TE. EuroQol-a new facility for the measurement of health-related quality of life. Health policy. 1990;16(3):199–208.

20. Devlin NJ, Shah KK, Feng Y, Mulhern B, Van Hout B. Valuing health-related quality of life: An EQ-5 D-5 L value set for England. Health economics. 2018;27(1):7–22.

21. Alshayban D, Joseph R. Health-related quality of life among patients with type 2 diabetes mellitus in Eastern Province, Saudi Arabia: A cross-sectional study. PLoS One. 2020;15(1):e0227573.

22. Khardali A, Kashan Syed N, Alqahtani SS, Qadri M, Meraya AM, Rajeh N, et al. Assessing medication adherence and their associated factors amongst type-2 diabetes mellitus patients of Jazan Province, Saudi Arabia: A single-center, cross-sectional study. Saudi Pharmaceutical Journal. 2024;32(1):1–8.

23. Shahabi N, Fakhri Y, Aghamolaei T, Hosseini Z, Homayuni A. Socio-personal factors affecting adherence to treatment in patients with type 2 diabetes: A systematic review and meta-analysis. Primary Care Diabetes. 2023;17(3):205–20.

24. Ahmed S, Saif-Ur-Rahman K, Dhungana RR, Ganbaatar G, Ashraf F, Yano Y, et al. Medication adherence and health-related quality of life among people with diabetes in bangladesh: A cross-sectional study. Endocrinology, Diabetes & Metabolism. 2023;6(5):e444.

25. Van Wilder L, Devleesschauwer B, Clays E, Pype P, Vandepitte S, De Smedt D. Polypharmacy and health-related quality of life/psychological distress among patients with chronic disease. Preventing Chronic Disease. 2022;19:E50.

26. Sabag A, Chang CR, Francois ME, Keating SE, Coombes JS, Johnson NA, et al. The effect of exercise on quality of life in type 2 diabetes: a systematic review and meta-analysis. Medicine & Science in Sports & Exercise. 2023;55(8):1353–65.

27. Park W, Lee D, editors. Efficacy of daily walking as a potential predictor of improved health-related quality of life in patients with type 2 diabetes in Korea. Healthcare; 2024: MDPI.

28. Mishra R, Sharma SK, Verma R, Kangra P, Dahiya P, Kumari P, et al. Medication adherence and quality of life among type-2 diabetes mellitus patients in India. World Journal of Diabetes. 2021;12(10):1740.

29. Martínez YV, Prado-Aguilar CA, Rascón-Pacheco RA, Valdivia-Martínez JJ. Quality of life associated with treatment adherence in patients with type 2 diabetes: a cross-sectional study. BMC health services research. 2008;8(1):164.

30. Azmi NL, Rosly NAM, Tang HC, Darof AFC, Zuki ND. Assessment of medication adherence and quality of life among patients with type 2 diabetes mellitus in a tertiary hospital in Kelantan, Malaysia. Journal of Pharmacy. 2021;1(2):79–86.

31. Highton PJ, Funnell MP, Gupta P, Zaccardi F, Lim L-L, Seidu S, et al. Improving medication adherence in type 2 diabetes: strategies for better clinical and economic outcomes. Diabetologia. 2026;69(3):541–56.

